# A Review of Population Pharmacokinetic Studies of Levetiracetam

**DOI:** 10.1101/2020.08.05.20167239

**Authors:** Zi-ran Li, Chen-yu Wang, Xiao Zhu, Zheng Jiao

**Author notes:** Corresponding author: Zheng Jiao, Professor, ^1^ Department of Pharmacy, Shanghai Chest Hospital, Shanghai Jiao Tong University 241 Huaihai West Road, 200030 Shanghai, P.R. China.

## Abstract

**Background:** Levetiracetam has been widely used as a treatment option for different types of epilepsy in both adults and children. Because of its large between-subject variability, several population pharmacokinetic studies have been performed to identify its pharmacokinetic covariates, and thus facilitate individualised therapy.

**Objective:** The aim of this review was to provide a synopsis for population pharmacokinetic studies of levetiracetam and explore the identified influencing covariates.

**Methods:** We systematically searched the PubMed and Embase databases from inception to 30 June, 2020. The information on study designs, target population, model characteristics, and identified covariates was summarised. Moreover, the pharmacokinetic profiles were compared among neonates, children, and adults.

**Results:** Fourteen studies were included, among which 2 involved neonates, 4 involved children, 2 involved both children and adults, and 6 involved only adults. The median value of apparent clearance for children (0.074 [range: 0.038–0.079] L/h/kg) was higher than that for adults (0.054 [range: 0.039–0.061] L/h/kg). Body weight was found to significantly influence the apparent clearance and volume of distribution significantly, whereas renal function influenced the clearance. Likewise, co-administration with enzyme-inducing antiepileptic drugs (such as carbamazepine and phenytoin) increased the drug clearance by 9%–22%, whereas co-administration with valproate acid decreased it by 18.8%.

**Conclusion:** Levetiracetam dose regimen is dependent on the body size and renal function of patients. Further studies are needed to evaluate levetiracetam pharmacokinetics in neonates and pregnant women.

**Key points:**

- This review identifies weight, renal function, daily dose, and postmenstrual age as the covariates that most likely influence the levetiracetam (LEV) pharmacokinetics.
- Children showed higher clearance per kilogram body weight than adults, indicating that a higher dosage is required for children per kilogram body weight.
- Further PPK studies are needed to evaluate LEV pharmacokinetics in special populations such as pregnant women and neonates.

## 1. Introduction

Levetiracetam (LEV) is a second-generation antiepileptic drug (AED) approved by the FDA to treat different types of epilepsy in both adults and children. Its mechanism of action is different to other AEDs. Studies have shown that LEV exerts its action on a new target, a synaptic vesicle protein 2A (SV2A), and inhibits neuronal (N)-type high-voltage-activated calcium channels [1, 2]. Because of its relatively safe adverse effect profile, LEV is recommended as a first-line therapy for patients with partial-onset seizures and it has replaced valproic acid (VPA) as the most frequently prescribed first-line antiepileptic drug in children since 2012 [3, 4].

LEV, after oral administration, is rapidly and almost completely absorbed (>95%). It reaches a peak plasma concentration (C_max_) in approximately 1 to 1.5 h after administration. LEV is not bound to the plasma protein, and its volume of distribution (V) ranges from 0.5 to 0.71 L/kg [5]. The drug is mainly metabolised by enzymatic hydrolysis of the acetamide group (27%), whereas the hepatic cytochrome P450 system plays only a small part (2.5%) [6]. Approximately, 66% of LEV is excreted unchanged by the kidney. Therefore, patients with renal impairment may need dose adjustments [6].

Moreover, LEV exhibits a high variability for children and pregnant women [7, 8]. Previous studies have shown lower LEV concentrations in children than in adults following the same dosage per kilogram bodyweight [7]. Likewise, enhanced LEV clearance has been reported in pregnant women [8]. Usually, in clinical practice, therapeutic drug monitoring (TDM) plays a crucial role in dose individualisation for these special patients [9]. However, the TDM approach has certain limitations, as it can only be implemented after treatment initiation. In such cases, population pharmacokinetic (PPK) analysis, which describes typical pharmacokinetic parameters of the target population and identifies factors that contribute to the variability of LEV, can help determining the appropriate initial LEV dosage for patients. Furthermore, through Bayesian forecasting, the PPK analysis can be developed as a powerful tool to estimate the individual PK parameters and develop dose individualisation, which is widely used in clinical practice [10, 11].

To date, several PPK studies on LEV have been conducted to identify the covariates that may have a significant effect on the pharmacokinetics. However, no study has summarised data concerning the PPK modelling of LEV. Thus, the aim of this review was to present an overview of these published PPK studies, summarise significant covariates affecting the LEV pharmacokinetics, and identify any knowledge gaps that remain to be explored.

## 2. Methods

### 2.1 Information Sources and Search Strategy

All PPK studies on LEV were systematically searched in the PubMed and Embase databases from inception until 30 June, 2020, as per the Systematic Reviews and Meta-Analyses (PRISMA) [12]. The pertinent PPK studies on LEV were identified using the following search terms: ‘levetiracetam’, ‘keppra’, or ‘elepsia’ and ‘population pharmacokinetic’, ‘pharmacokinetic modeling’, ‘nonlinear mixed effect model’, ‘NONMEM’, ‘Pmetrics’, ‘WINNONMIX’, ‘ADAPT’, ‘P-PHARM’, ‘nlmixed’, ‘NLME’, ‘USC*PACK’, or ‘MONOLIX’. In addition, the reference lists of the selected articles were also checked to identify any related studies. Two authors independently performed the literature search. A third senior investigator was consulted to resolve any discrepancies.

All studies identified from databases and other sources were screened to assess their eligibility. A study was considered eligible for inclusion in this review if it met the following criteria: (1) the study population included healthy volunteers or patients; (2) LEV was used as the study drug, regardless of its formulation; (3) the study was focused on PPKs or pharmacokinetic/pharmacodynamic (PK/PD) analysis. A publication was excluded if: (1) it was a review or only focused on the methodology, algorithm, or software considerations; (2) it was published in a non-English language; and (3) the information on methodology or pharmacokinetics was insufficient.

### 2.2 Data Extraction

The following information was extracted from the included articles: (1) the characteristics of the target population (patients or healthy subjects) and their demographics (e.g. age, weight range, and sex); (2) the study design (e.g. type of study, number of participants and collected samples, sampling design, dosage regimens, and LEV formulations); and (3) the information on PPK analyses such as data analysis software, structural models, between-subject (BSV) variability and residual unexplained variability (RUV), parameter estimates, covariates, and model evaluation approaches.

### 2.3 Comparison of Studies

Patient characteristics, population analysis strategies, pharmacokinetic parameters, and the screened covariates of each study were summarised in a tabular format. The concentration-time profiles of neonates (3 kg, postmenstrual age [PMA] 40 weeks), children (30 kg, 10 years), and adults (70 kg, 40 years), for 20 mg/kg LEV were plotted according to the established PPK model and study cohorts in each study. The serum creatinine (Cr) was set to 0.5 mg/dL for neonates and the estimated glomerular filtration rate (eGFR) was set to 90 mL/min/1.73 m^2^ for children and adults. The patients were assumed to have received monotherapy and reached a steady state. The sex was set to male.

The effect of significant covariates for clearance (CL) in each study was summarised using a forest plot, in which, a less than 20% change in CL was not regarded to have clinical relevance. The effect of each covariate on CL is displayed by the ratio of CL in the range of each covariate dividing the typical CL value in each study. For binary covariates such as sex, 0 and 1 were used. For continuous covariates included only in one model, we used their minimum and maximum values in the model. For continuous covariates included in several models, we scaled them to the same range for comparison as follows: weight was scaled into three groups including neonates (1–10 kg), children (16–40 kg), and adults (40–100 kg). The range of eGFR was set as 30–100 mL/min/1.73 m^2^. The daily dose of LEV was set as 500–3000 mg.

## 3. Results

### 3.1 Study Identification

A total of 143 and 79 studies were identified from the PubMed and Embase database searches, respectively. One additional study was identified via the reference list of the selected articles. After the removal of duplicates, 184 studies were screened. Several screened studies were then disqualified, according to the aforementioned exclusion criteria. Eighteen full-text articles were assessed for eligibility. Among these, three were excluded because of missing PPK parameter estimates, and one was excluded because its data were reported in other articles. Finally, 14 studies were included in the review for further analysis. The PRISMA diagram of study identification is presented in Fig. 1.

**Fig. 1.**
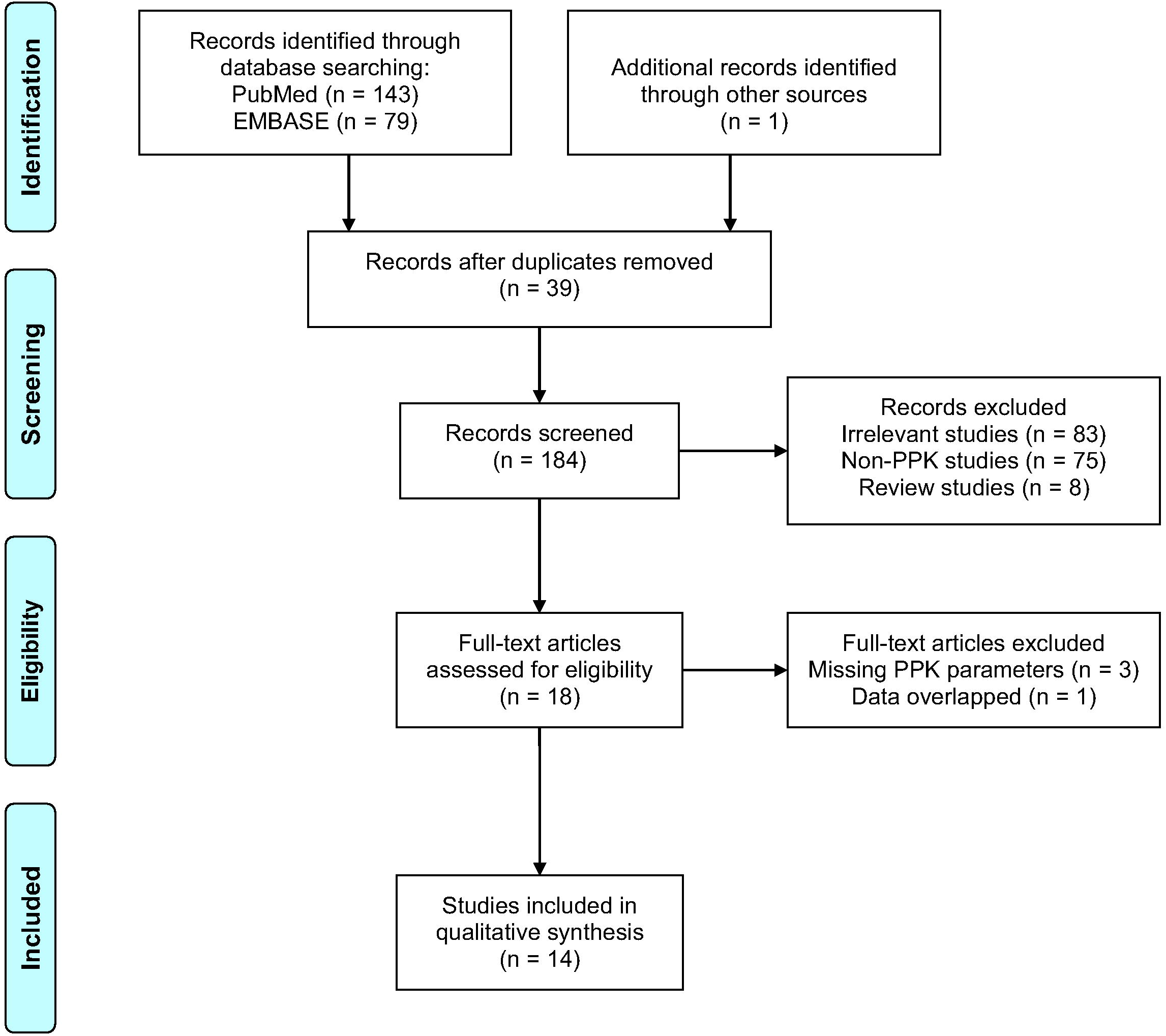
PRISMA flow diagram for the identification of LEV PPK studies.

### 3.2 Study Characteristics

All included studies were published between 2007 and 2020. The characteristics of each study are summarised in Table 1. Sneck et al. [13] enrolled only healthy subjects, Pigeolet et al. included both patients and healthy subjects [14], and the other studies were conducted in patients with epilepsy [15-26]. Eight of the fourteen studies enrolled paediatric patients [15, 17, 18, 20, 21, 24-26]; 2 studies were conducted in neonates [18, 21], 4 studies in children less than 18 years of age [15, 20, 25, 26], and 2 studies included both children and adults [17, 24]. Three study only enrolled adults [13, 14, 19], and the other three studies included elderly patients of over 70 years [16, 22, 23].

**Table 1.**
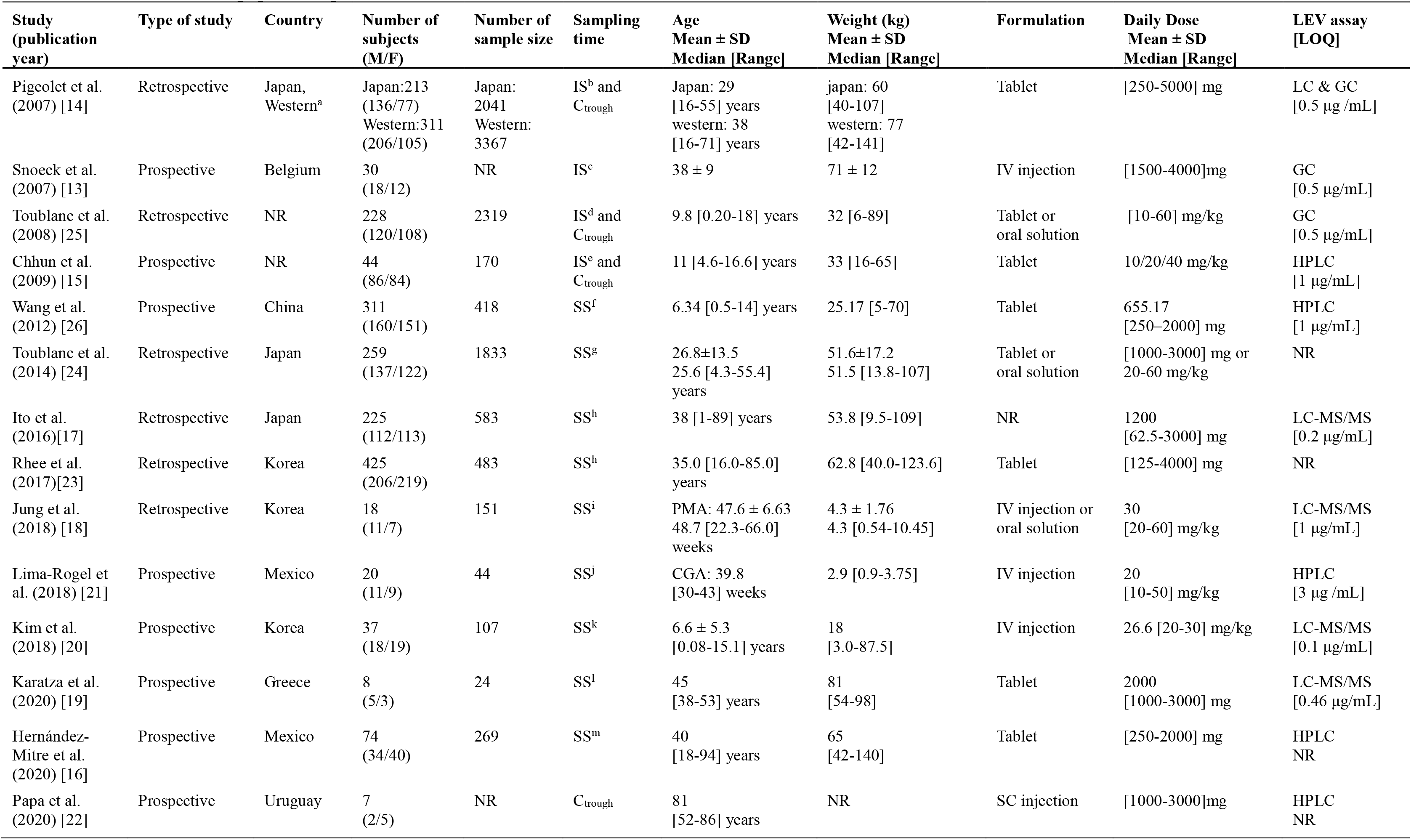
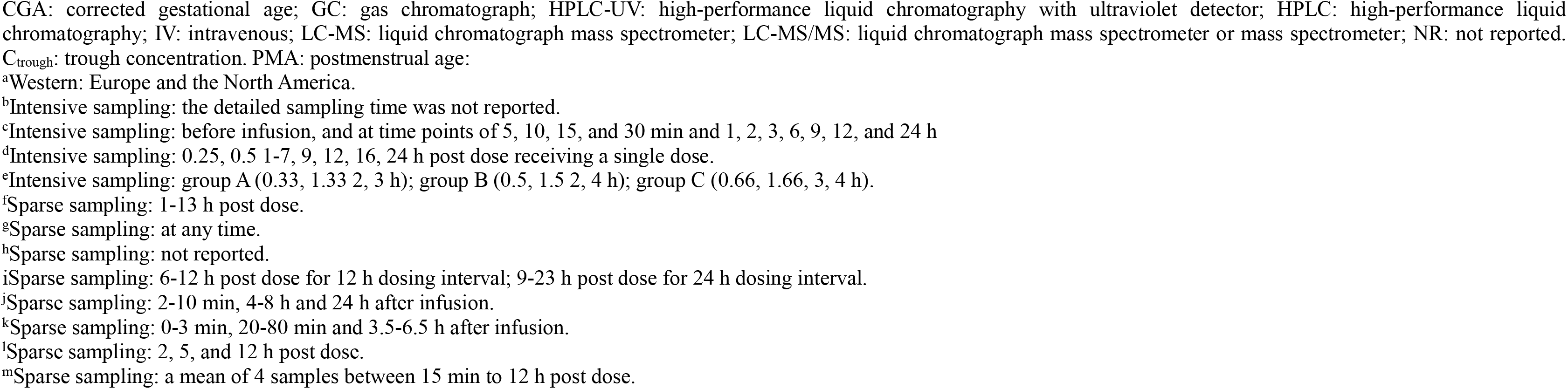
Characteristics of population pharmacokinetic studies included in the systematic review.

The PPK model developed by Karatza et al. [19] and Papa et al. [22] were built using Monolix software and other models were built using NONMEM software. The LEV PK for oral formulations was described with a one-compartment model with first-order absorption and elimination, and for injections was described with a one- or two- compartment model with first order elimination. Two studies conducted in neonates [18, 21], three in children [20, 26], and five in adults were based on sparse sampling data from the routine clinical TDM settings [16, 17, 19, 22, 23], and other four studies included intensive sampling data [13-15, 25]. The number of participants in each study ranged from 7 to 524, with LEV observations per individual ranging from 1 to 10. The LEV doses ranged between 4.5 and 130 mg/kg. High-performance liquid chromatography, high-performance liquid chromatography with tandem mass spectrometry, and gas chromatography were employed as LEV bioassay methods. The lowest limit for quantitative assay ranged between 0.1 and 3 μg/mL.

### 3.3 Comparison of Studies

Two PPK studies were conducted in neonates [18, 21]. For neonates (PMA 40 weeks, 3 kg), the CL estimated by Jung et al. [18] was 0.043 L/h/kg, similar to 0.049 L/h/kg estimated by Lima-Rogel et al. [21]. However, the V estimated by Jung et al. [18] was much higher than that by Lima-Rogel et al. [21] (1.07 vs 0.65 L/h/kg).

Most studies conducted in children and adults displayed similar concentration-time profiles, as shown in Fig. 2b and 2c. However, the model developed by Wang et al. [26] showed a considerably higher trough concentration (C_trough_) than the others, and this could be explained by the fact that the CL estimated by Wang et al. [26] was considerably lower than that estimated in the other studies conducted in children (30 kg, 10 years) (0.038 vs 0.067–0.079 L/h/kg) [17-19, 21].

**Fig. 2.**
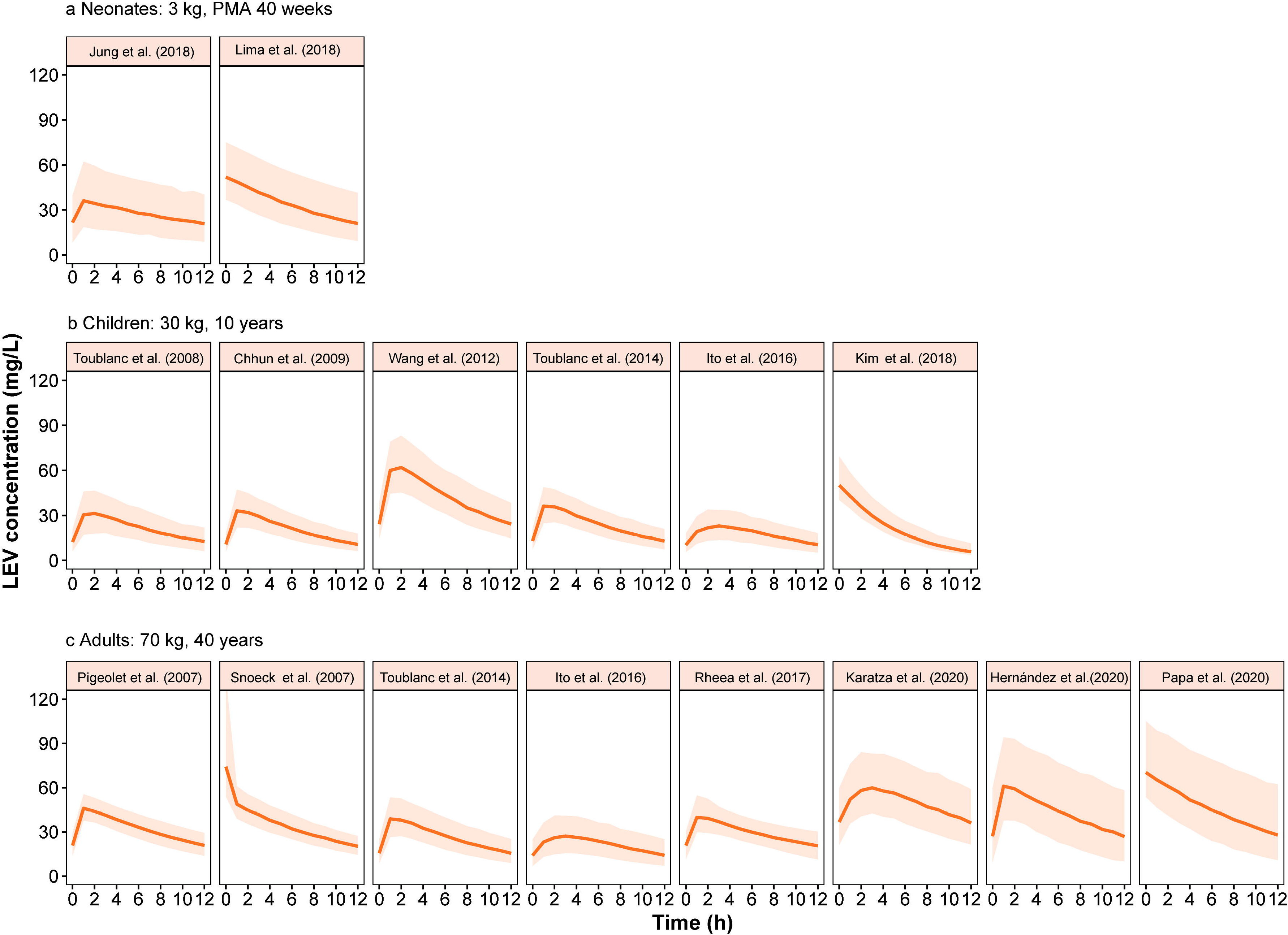
Concentration-time profiles of LEV at steady state for neonates (a), children (b), and adults (c) in retrieved studies. The solid line represents median of the simulated concentration-time profile. The light shadows represent the 10^th^-90^th^ percentiles of the simulated concentration-time profiles. The estimated glomerular filtration rate (eGFR) was set to 90 mL/min/1.73 m^2^ for children and adults. The creatinine (Cr) for neonates was set to 0.5 mg/dL. All patients were assumed to be male receiving LEV monotherapy at a dose of 20 mg/kg. PMA: postmenstrual age.

For neonates (3 kg, PMA 40 weeks), children (30 kg, 10 years), and adults (70 kg, 40 years), the estimated median (range) of CL was: 0.046 (0.043–0.049) L/h/kg for neonates, 0.074 (0.038–0.079) L/h/kg for children, and 0.054 (0.039–0.061) L/h/kg for adults. The median CL in neonates and adults was lower than that in children. On the contrary, the V was determined as: 0.86 (0.65–1.07) L/kg for neonates, 0.65 (0.40–0.75) L/kg for children, and 0.60 (0.42–0.93) L/kg for adults. The V for neonates was higher than that for children and adults.

The aim of almost all included PPK studies was to identify the potential covariates to describe the BSV of LEV pharmacokinetics, except for that of Papa et al. [22], in which no covariates were investigated because of limited patient numbers. The covariate screening process is summarised in Table 2. All covariates that were investigated and retained in the final model are summarised in Fig. 3. The most frequently screened covariates included weight, eGFR, co-administered medication, sex, and age. The covariates identified for CL included weight, eGFR, co-administered medication, daily dose, sex, Cr, and PMA.

**Table 2.**
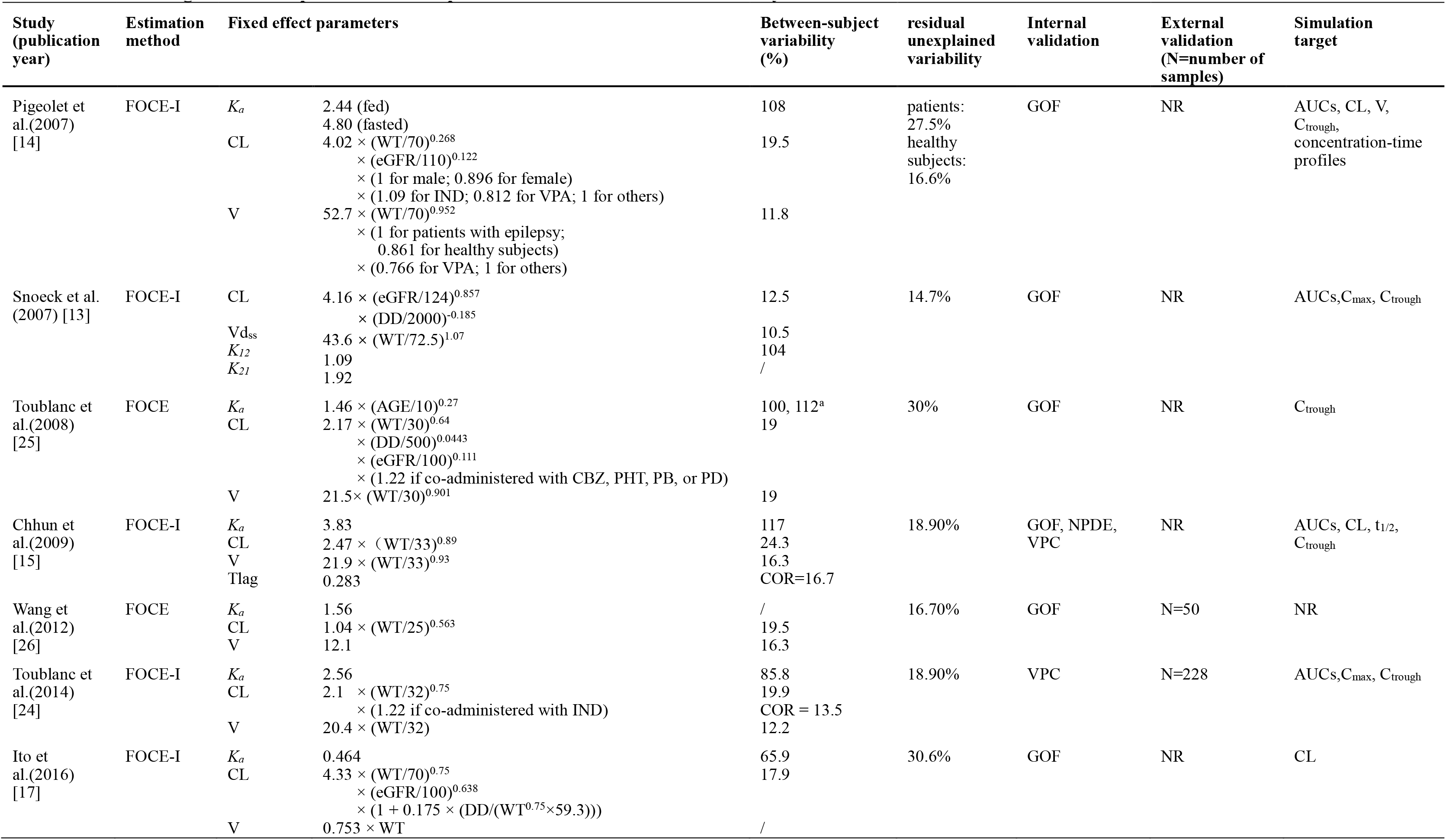
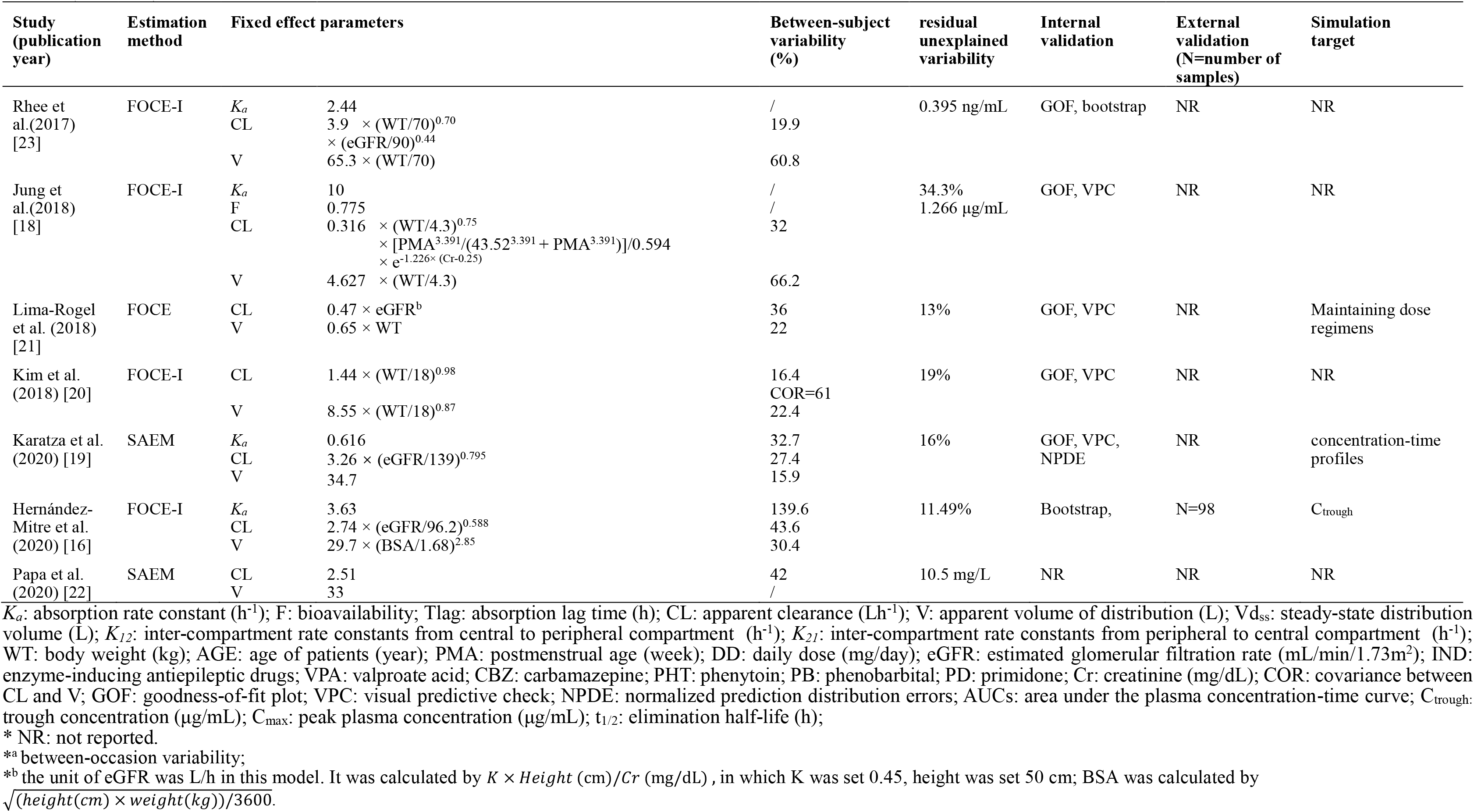
Model strategies and final pharmacokinetic parameters of included studies in the systematic review.

**Fig. 3.**
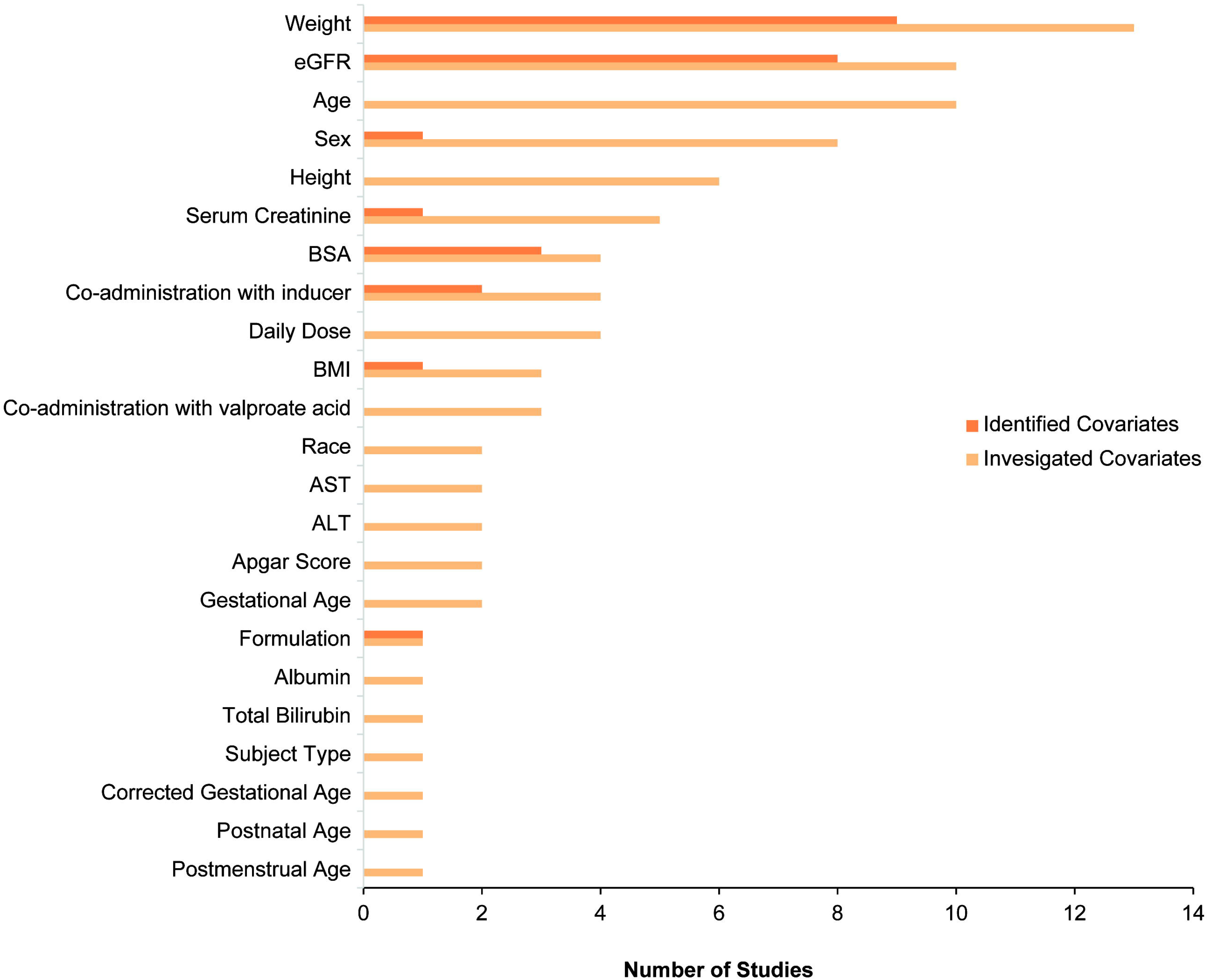
Investigated and identified covariates for clearance of LEV. eGFR: the estimated glomerular filtration rate; BSA: body surface area; IND: co517 administration with enzyme-inducing antiepileptic drugs; AST: aspartate aminotransferase; ALT: alanine aminotransferase; BMI: body mass index; GA: gestational age; VPA: co-administration with valproate acid; CGA: corrected gestational age; PNA: postnatal age; PMA: postmenstrual age.

**Table 3.** List of tested and significant covariates in the models.

| Study<br>(publication<br>year) | Tested covariates |  |  | Covariate selection criteria |  | Significant covariates |  |  |
| --- | --- | --- | --- | --- | --- | --- | --- | --- |
| | Demographic | Laboratory examination | Co-administration | Forward<br>inclusion | Backward<br>elimination | $K_a$ | CL | V |
| Pigeolet et al.<br>(2007) [14] | Weight, Height, BMI, BSA,<br>Sex, Race, Subject Type<sup>a</sup> | eGFR, Serum Creatinine | AEDs, Benzodiazepine | $P < 0.05$ | $P < 0.005$ | NR | Weight, eGFR, Sex,<br>IND, VPA | Weight, Subject<br>Type, VPA |
| Snoeck et al.<br>(2007) [13] | Age, Weight, Sex, Daily Dose | eGFR | NR | $P < 0.05$ | $P < 0.005$ | NR | eGFR, Daily Dose | Weight |
| Toublanc et al.<br>(2008)<br>[25] | Age, Weight, BSA, BMI, Sex,<br>Race, Daily Dose | eGFR | Inducer, Inhibitor, Benzodiazepine | $P < 0.05$ | $P < 0.005$ | Age | Weight, eGFR,<br>Daily Dose, IND | Weight |
| Chhun et al.<br>(2009) [15] | Age, Weight, Sex | eGFR | NR | $P < 0.05$ | $P < 0.01$ | NR | Weight | Weight |
| Wang et al.<br>(2012) [26] | Age, Weight, Daily Dose | NR | VPA, LTG, CBZ, OXC, TPM | $P < 0.005$ | NR | NR | Weight | NR |
| Toublanc et al.<br>(2014)<br>[24] | Age, Weight, Formulation | NR | Neutral, Inducer, Inhibitor, A<br>combination of inducer and inhibitor | $P < 0.05$ | $P < 0.001$ | NR | Weight, IND | Weight |
| Ito et al.<br>(2016) [17] | Age, Weight, Sex, Daily Dose | eGFR, ALT, AST | AEDs | $P < 0.01$ | $P < 0.005$ | NR | Weight, eGFR,<br>Daily Dose | Weight |
| Rhee et al.<br>(2017) [23] | Age, Weight, Height, Sex | eGFR, Serum Creatinine,<br>AST, ALT, Albumin,<br>Total bilirubin | VPA, LTG, CBZ, OXC, TPM,<br>Phenytoin, Phenobarbital, Pregabalin,<br>Zonisamide, Clobazam | $P < 0.05$ | $P < 0.001$ | NR | Weight, eGFR | Weight |
| Jung et al.<br>(2018) [18] | PMA, GA, Weight, Apg1,<br>Apg5 | Serum Creatinine | NR | $P < 0.01$ | $P < 0.001$ | NR | Weight, PMA,<br>Creatinine | Weight |
| Lima-Rogel et<br>al. (2018) [21] | PNA, GA, CGA, Weight,<br>Height, BSA, Apgar Score,<br>Sex | eGFR, Serum Creatinine | NR | $P < 0.05$ | $P < 0.001$ | NR | eGFR | Weight |
| Kim et al.<br>(2018) [20] | Age, Weight, Height, Sex | eGFR | NR | $P < 0.05$ | NR | NR | Weight | Weight |
| Karatza et al.<br>(2020) [19] | Age, Weight, Height, Daily<br>Dose | eGFR | NR | $P < 0.01$ | NR | NR | eGFR | NR |
| Hernández-Mitre et al.<br>(2020) [16] | Age, Weight, Height, BSA,<br>BMI | eGFR, Serum Creatinine | Inducer, Inhibitor | $P < 0.05$ | $P < 0.01$ | NR | eGFR | BSA |
| Papa et al.<br>(2020) [22] | NR | NR | NR | NR | NR | NR | NR | NR |

BMI: body mass index; BSA: body surface area; CGA: corrected gestational age; PMA: postmenstrual age; PNA: postnatal age; GA: gestational age; ALT: alanine aminotransferase; AST: aspartate aminotransferase; eGFR: estimated glomerular filtration rate; IND: enzyme-inducing antiepileptic drugs; AEDs: antiepileptic drugs; VPA: valproate acid; LTG: lamotrigine; CBZ: carbamazepine; OXC: oxcarbazepine; TPM: topiramate; Apg1: Apgar score = 1; Apg5: Apgar score = 5; NR: not reported. Inducer: enzyme-inducing AEDs as carbamazepine, oxcarbazepine, primidone, phenytoin and phenobarbital; Inhibitor: enzyme-inhibiting AEDs as valproate acid;
<sup>a</sup> Subject Type: Healthy subject or patients with epilepsy;

Nine of the fourteen studies indicated that weight is associated with the CL of LEV [14, 15, 17, 18, 20, 23-26]. Four studies [13, 16, 19, 21], in contrast, found no relationship between weight and CL, but an association between eGFR and CL. Weight had a significant influence on CL in all three age groups (Fig. 4). Moreover, the eGFR could explain the BSV of CL in eight studies [13, 14, 16, 17, 19, 21, 23, 25], five of which showed the clinical significance of an influence on CL larger than ± 20% [13, 16, 17, 19, 23]. According to these studies, in patients with renal impairment (eGFR: 30 mL/min/1.73 m^2^), the CL was 44% (12.5%-70.5%) lower than that in patients with median renal function in each study.

**Fig. 4.**
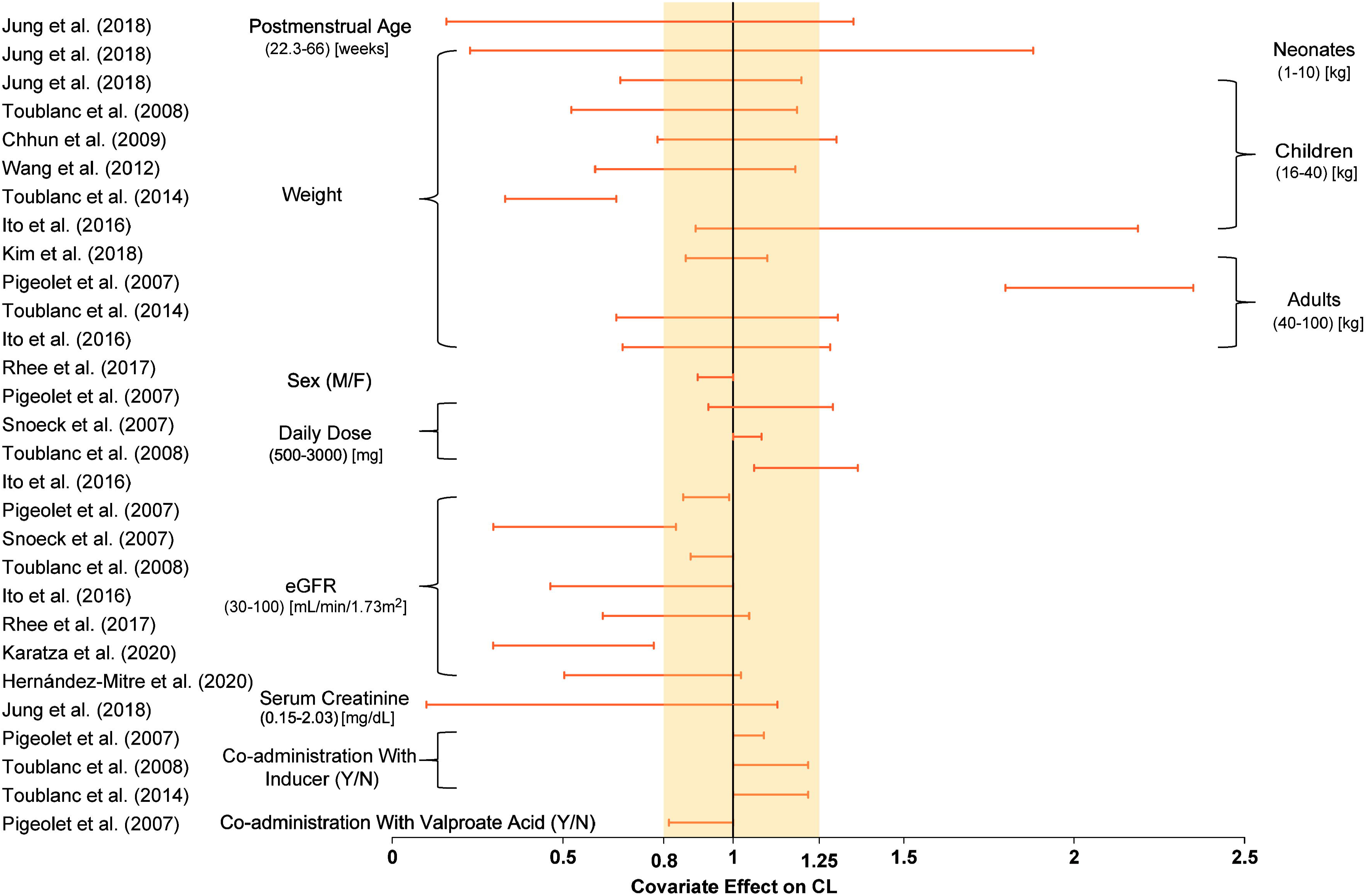
Covariate effect on the clearance of LEV. The horizontal bars represent the covariate effect on clearance in each study. The typical value of clearance in each study was considered to be 1. The effect of each covariate for clearance is displayed by the ratio of clearance in the range of each covariate to the typical clearance value. The shaded gray area ranges from 0.8 to 1.25. eGFR: the estimated glomerular filtration rate; IND: co-administration with enzyme-inducing antiepileptic drugs; VPA: co-administration with valproate acid; PMA: postmenstrual age; Y: yes, co-administration; N: no, not co-administration; M: male; F: female.

Three studies reported that the co-administration of enzyme-inducing antiepileptic drugs, such as carbamazepine and phenobarbital, increased the CL by 9%–22%, [14, 24, 25], whereas co-administration with VPA decreased it by approximately 18.8% [14]. However, these co-medications may not have any clinical significance, as the change in CL was found to be less than 22%. Three studies identified that the daily dose could influence the CL [13, 14, 25]. One study showed that females had 10.4% lower CL than males [14]. For neonates, the PMA and Cr were found to significantly affect the CL in one study [18].

In all studies included, the BSV was described by exponential models. The median (range) of BSV is as follows: CL, 19.9% (12.5%–43.6%) [n = 14]; V, 17.6% (10.5%–66.2%) [n = 12]; and Ka, 100% (32.7%–117%) [n = 7]. The RUV was commonly described by the proportional models to range from 11.49% to 34.3%; a higher RUV was reported by the study in neonates than in other studies [18]. One study showed that the RUV in patients with epilepsy was considerably higher than that in healthy subjects (27.5% and 16.6%, respectively) [14].

All models were internally evaluated. The most frequently used method was goodness-of-fit plots, which showed the closeness between prediction and observation. Only three studies performed external evaluation and showed acceptable predictability [16, 24, 26].

Nine studies performed a model-based simulation to display the effects of covariates and propose dosing regimens [13-17, 19, 21, 24, 25]. The influence of age, weight, race, and eGFR on LEV clearance, area under the concentration-time curve, C_trough_, and C_max_ were investigated. Weight and eGFR were found to have significant influences on dose recommendation. For children, Toublanc et al. (2008) [25] recommended that the dosage should be differentiated as per body weight: 10 mg/kg for children less than 20 kg; 250 mg twice daily for children between 20 and 40 kg, and 500 mg twice daily for children more than 40 kg. For adults, Hernández-Mitre et al. [16] recommended to adjust the dosage according to eGFR to achieve the trough LEV concentrations between 12 and 46 μg/mL.

## 4. Discussion

LEV is one of the most widely used AEDs. There has been a continued interest in studying the LEV pharmacokinetics from the past few decades, and several PPK studies have been reported to identify the sources of variability. This is the first review to summarise the knowledge concerning the population pharmacokinetic modelling of LEV.

All studies conducted in adult patients showed similar concentration-time profiles for patients with the same demographic characteristics. Moreover, two population studies investigated the ethnic variance for LEV, and found no obvious difference [14, 24]. Toublanc et al. (2014) [24] developed a PPK model based on Japanese patients and extrapolated it to North American patients through an external evaluation; the model showed satisfactory predictive performance. Pigeolet et al. [14] enrolled Japanese, European, and North American patients to conduct a PPK analysis and did not find any significant difference among these sub-populations. However, Wang et al. [26] showed a lower clearance in Chinese children than in Japanese, European, and North American sub-population. This difference warrants further investigation.

The pharmacokinetics of LEV showed large differences between children and adults. The children showed a higher CL and a lower V per kilogram body weight than the adults with the same dosage per kilogram bodyweight. The difference in CL between children and adults may be attributed to the decrease in blood flow, increase in body fat/lean mass ratio, and decrease in total body water owing to ageing. One study reported that the CL of LEV rapidly increased after birth and decreased during ageing, which is similar to the growth tendency of renal function [27]. The renal function increases rapidly during the first 2 to 3 years of life and then decreases with ageing in proportion to the physiological function decrease [28]. As LEV is mainly eliminated by the kidney, and children showed a higher renal function than adults, the alternation in renal function during ageing may account for the difference in CL between children and adults [29, 30]. Thus, a higher dosage per weight is required in children than in adults for the same trough concentration.

As LEV is mainly eliminated by the kidney (66%), renal function indicators, such as eGFR and Cr, may influence the CL of LEV drastically [6]. Karatza et al. [19] found that the CL of LEV decreased by approximately 27.6% for patients with renal insufficiency (60 mL/min/1.73 m^2^), and 58.3% in patients with renal impairment (30 mL/min/1.73 m^2^) compared with individuals with a normal renal function (90 mL/min/1.73 m^2^). Jung et al. [18] found that the CL of LEV may decrease by approximately 37% in neonates, with an increase in Cr by 30%. Consequently, the dosing regimen for patients with renal impairment needs to be adjusted.

Three studies identified that the daily dose affected the CL of LEV, which may indicate a non-linear elimination of LEV [13, 17, 25]. Toublanc et al. (2008) [25] and Ito et al. [17] found higher dosage increased the CL. On the contrary, Snoeck et al. found higher dosage could decrease the CL [13]. This influence may result from the dose adjustment in clinical settings, where a higher dosage was adjusted for patients with a higher CL [31]. Further exploration of the potential non-linear PK of LEV is still needed.

Only 2.5% of LEV is metabolised by the hepatic cytochrome P450 system [6]. Therefore, the CL of LEV may be less affected by the CYP enzyme inducers or inhibitors. Yet, three PPK studies showed that co-administration with enzyme-inducing antiepileptic drugs, such as carbamazepine and phenobarbital, could increase the CL of LEV by 9%–22% [14, 24, 25]. The possible enzyme induction effect requires further investigation.

Moreover, Pigeolet et al. [14] found that the co-administration of LEV and VPA decreased the CL of LEV by 18.8% in their PPK analysis. This finding was consistent with the observations of Perucca et al. and May et al. [7, 32] that VPA could influence the CL of LEV in classical PK studies. However, Coupez et al. [33] found that VPA could not alter the pharmacokinetics of LEV during *in vitro* and *in vivo* studies. VPA is the inhibitor of epoxide hydrolase and the CYP2C isoenzymes, none of which are involved in the pharmacokinetics of LEV. Thus, the mechanism of VPA and LEV interaction remains to be clarified in future studies.

Ten studies reported that the V has a significant relationship with weight [13-15, 17, 18, 20, 21, 23-25], except three studies fixed it to be 12.5, 34.7, and 33L, respectively [19, 22, 26] and one study found that the V has a significant relationship with body surface area (BSA) [16]. Pigeolet et al. [14] found that the V was 13.9% lower in healthy subjects than in patients with epilepsy. Moreover, in this study, V was found to decrease by 23.4% in patients co-administered with VPA. However, its mechanism remains uncertain.

Four studies reported a PK/PD analysis and their findings were not consistent [15, 19, 20, 34]. Chhun et al. [15] performed the exposure-response analysis for LEV, wherein the patients were classified into two groups (seizure-free for at least 3 months or not). The C_trough_ showed no difference between the groups. In the other study, Snoeck et al. [34] utilised a model to predict the reduction in seizure frequency with PK of LEV, and indicated a clear relationship between LEV dose and effect. Kim et al. [20] found Weibull distribution could best describe the time to seizure recurrence with LEV administration, but no significant covariates were identified. Karatza et al. [19] found a strong PK/PD linkage between LEV concentrations and seizure recurrence. In this study, patients with a higher daily dose and higher eGFR were more likely to have a seizure recurrence, but only eight patients were recruited [19]. Thus, further prospective PK/PD studies of LEV with larger samples should be conducted to clarify the exposure-response relationship.

Owing to the linear pharmacokinetics, low protein binding, and better safety profile of LEV, it has been increasingly used off-label in the treatment of neonatal seizures [35]. Accumulating evidence on the safety and efficacy of LEV in neonatal seizures support its use as a second-line therapy [36]. So far, two PPK studies have been conducted in neonates, using either an oral solution or IV injection formulation [18, 21]. The CL for neonates has been found to be highly influenced by Cr, which should be considered for dose adjustment for neonates. Besides, the RUV in models for neonates was the highest among all the included studies. The large differences between neonates may be owing to the physiological differences such as total body water-to-fat ratios, gastrointestinal motility, drug-metabolising enzyme activity, and renal function [37, 38]. Further studies are still needed to identify influential factors and describe the variability.

LEV shows a good foetal safety profile and has been widely used for pregnant women with epilepsy [39-42]. However, the CL of LEV is higher in pregnant women than in non-pregnant individuals, which could lead to the failure of seizure control [8, 43]. Thus, the TDM of LEV in pregnant patients is necessary to maintain the appropriate concentration [44]. However, the data reported for LEV in pregnant women are insufficient. As PPK modelling can help investigate the pharmacokinetic characteristics in special populations from sparse samples, it offers an appropriate approach for dose individualisation in pregnant women, and it has been used for individualised lamotrigine therapy in pregnant women [45]. Thus, further PPK studies are required to characterise LEV pharmacokinetics in pregnant women in the future.

## 5. Conclusions

The PPK studies of LEV were systematically reviewed. The pharmacokinetics of LEV differed between children and adults, such that children show a higher CL and a lower V than adults following the same dosage per kilogram body weight. The review establishes that LEV dose individualisation should be dependent on the body size and renal function of patients. Further PPK studies are required in neonates and pregnant women to establish the LEV pharmacokinetic profile.

## Data Availability

No Data

## Declarations

### Ethics approval

Not applicable.

### Consent to participate

Not applicable.

### Consent for publication

Not applicable.

## Availability of data and material

### Code availability

Not applicable.

### Author contributions

Zi-ran Li, Chen-yu Wang, and Zheng Jiao designed the review and planned the work that led to the manuscript. Zi-ran Li and Chen-yu Wang performed the literature search and data analysis. Zi-ran Li, Xiao Zhu, and Zheng Jiao drafted and revised the manuscript. All authors approved the final version of this manuscript.

## Acknowledgments

The authors would like to sincerely thank *Dr. Silvia Romano Moreno* from Facultad de Ciencias Químicas, Universidad Autónoma de San Luis Potosí, San Luis Potosí, México; *Dr. V. Lima-Rogel* from the Neonatal Intensive Care Unit, Hospital Central “Dr. Ignacio Morones Prieto”, San Luis Potosí, México; *Dr. Yun Seob Jung* from the Department of Pharmacology, Yonsei University College of Medicine, Korea; and *Dr. Brigitte Lacroix* from UCB Pharma SA, Belgium for providing details about the research and active discussions on the coding. We would like to thank Hai-ni Wen MPharm; Yi-wei Yin, Pharm. D from Shanghai Chest Hospital; and PhD candidate Xiao-qin Liu for their critical comments. We would also like to thank Editage (www.editage.cn) for English language editing.

## Reference

1. Niespodziany I, Klitgaard H, Margineanu DG. Levetiracetam inhibits the high-voltage-activated Ca(2+) current in pyramidal neurones of rat hippocampal slices. Neurosci Lett. 2001;306(1-2):5–8.

2. Lukyanetz EA, Shkryl VM, Kostyuk PG. Selective blockade of N-type calcium channels by levetiracetam. Epilepsia. 2002;43(1):9–18.

3. Weijenberg A, Bos JHJ, Schuiling-Veninga CCM, Brouwer OF, Callenbach PMC. Antiepileptic drug prescription in Dutch children from 2006-2014 using pharmacy-dispensing data. Epilepsy Res. 2018;146:21–7.

4. Glauser T, Ben-Menachem E, Bourgeois B, Cnaan A, Guerreiro C, Kälviäinen R, et al. Updated ILAE evidence review of antiepileptic drug efficacy and effectiveness as initial monotherapy for epileptic seizures and syndromes. Epilepsia. 2013;54(3):551–63.

5. Patsalos PN. Clinical pharmacokinetics of levetiracetam. Clin Pharmacokinet. 2004;43(11):707–24.

6. Strolin Benedetti M, Whomsley R, Nicolas J-M, Young C, Baltes E. Pharmacokinetics and metabolism of 14C-levetiracetam, a new antiepileptic agent, in healthy volunteers. Eur J Clin Pharmacol. 2003;59(8-9):621–30.

7. May TW, Rambeck B, Jürgens U. Serum concentrations of Levetiracetam in epileptic patients: the influence of dose and co-medication. Ther Drug Monit. 2003;25(6):690–9.

8. Reisinger TL, Newman M, Loring DW, Pennell PB, Meador KJ. Antiepileptic drug clearance and seizure frequency during pregnancy in women with epilepsy. Epilepsy Behav. 2013;29(1):13–8.

9. Naik GS, Kodagali R, Mathew BS, Thomas M, Prabha R, Mathew V, et al. Therapeutic Drug Monitoring of Levetiracetam and Lamotrigine: Is There a Need? Ther Drug Monit. 2015;37(4):437–44.

10. Sheiner LB, Rosenberg B, Melmon KL. Modelling of individual pharmacokinetics for computer-aided drug dosage. Comput Biomed Res. 1972;5(5):411–59.

11. Sasaki T, Tabuchi H, Higuchi S, Ieiri I. Warfarin-dosing algorithm based on a population pharmacokinetic/pharmacodynamic model combined with Bayesian forecasting. Pharmacogenomics. 2009;10(8):1257–66.

12. Shamseer L, Moher D, Clarke M, Ghersi D, Liberati A, Petticrew M, et al. Preferred reporting items for systematic review and meta-analysis protocols (PRISMA-P) 2015: elaboration and explanation. BMJ. 2015;349:g7647.

13. Snoeck E, Jacqmin P, Sargentini-Maier ML, Stockis A. Modeling and simulation of intravenous levetiracetam pharmacokinetic profiles in children to evaluate dose adaptation rules. Epilepsy Res. 2007;76(2-3):140–7.

14. Pigeolet E, Jacqmin P, Sargentini-Maier ML, Stockis A. Population pharmacokinetics of levetiracetam in Japanese and Western adults. Clin Pharmacokinet. 2007;46(6):503–12.

15. Chhun S, Jullien V, Rey E, Dulac O, Chiron C, Pons G. Population pharmacokinetics of levetiracetam and dosing recommendation in children with epilepsy. Epilepsia. 2009;50(5):1150–7.

16. Hernández-Mitre MP, Medellín-Garibay SE, Rodríguez-Leyva I, Rodríguez-Pinal CJ, Zarazúa S, Jung-Cook HH, et al. Population Pharmacokinetics and Dosing Recommendations of Levetiracetam in Adult and Elderly Patients With Epilepsy. J Pharm Sci. 2020;109(6):2070–8.

17. Ito S, Yano I, Hashi S, Tsuda M, Sugimoto M, Yonezawa A, et al. Population Pharmacokinetic Modeling of Levetiracetam in Pediatric and Adult Patients With Epilepsy by Using Routinely Monitored Data. Ther Drug Monit. 2016;38(3):371–8.

18. Jung YS, Lee SM, Park MS, Park K. Population pharmacokinetic model of levetiracetam in Korean neonates with seizures. Int J Clin Pharmacol Ther. 2018;56(5):217–23.

19. Karatza E, Markantonis SL, Savvidou A, Verentzioti A, Siatouni A, Alexoudi A, et al. Pharmacokinetic and pharmacodynamic modeling of levetiracetam: investigation of factors affecting the clinical outcome. Xenobiotica. 2020;50(9):1090–1100.

20. Kim MJ, Yum MS, Yeh HR, Ko TS, Lim HS. Pharmacokinetic and Pharmacodynamic Evaluation of Intravenous Levetiracetam in Children With Epilepsy. J Clin Pharmacol. 2018;58(12):1586–96.

21. Lima-Rogel V, López-López EJ, Medellín-Garibay SE, Gómez-Ruiz LM, Romero-Méndez C, Milán-Segovia RC, et al. Population pharmacokinetics of levetiracetam in neonates with seizures. J Clin Pharm Ther. 2018;43(3):422–9.

22. Papa P, Oricchio F, Ginés M, Maldonado C, Tashjian A, Ibarra M, et al. Pharmacokinetics of Subcutaneous Levetiracetam in Palliative Care Patients. J Palliat Med. 2020;10.1089

23. Rhee SJ, Shin JW, Lee S, Moon J, Kim TJ, Jung KY, et al. Population pharmacokinetics and dose-response relationship of levetiracetam in adult patients with epilepsy. Epilepsy Res. 2017;132:8–14.

24. Toublanc N, Lacroix BD, Yamamoto J. Development of an integrated population pharmacokinetic model for oral levetiracetam in populations of various ages and ethnicities. Drug Metab Pharmacokinet. 2014;29(1):61–8.

25. Toublanc N, Sargentini-Maier ML, Lacroix B, Jacqmin P, Stockis A. Retrospective population pharmacokinetic analysis of levetiracetam in children and adolescents with epilepsy: dosing recommendations. Clin Pharmacokinet. 2008;47(5):333–41.

26. Wang YH, Wang L, Lu W, Shang DW, Wei MJ, Wu Y. Population pharmacokinetics modeling of levetiracetam in Chinese children with epilepsy. Acta Pharmacol Sin. 2012;33(6):845–51.

27. Italiano D, Perucca E. Clinical pharmacokinetics of new-generation antiepileptic drugs at the extremes of age: an update. Clin Pharmacokinet. 2013;52(8):627–45.

28. Hämmerlein A, Derendorf H, Lowenthal DT. Pharmacokinetic and pharmacodynamic changes in the elderly. Clinical implications. Clin Pharmacokinet. 1998;35(1):49–64.

29. Dahlin MG, Wide K, Ohman I. Age and comedications influence levetiracetam pharmacokinetics in children. Pediatr Neurol. 2010;43(4):231–5.

30. Johannessen Landmark C, Baftiu A, Tysse I, Valsø B, Larsson PG, Rytter E, et al. Pharmacokinetic variability of four newer antiepileptic drugs, lamotrigine, levetiracetam, oxcarbazepine, and topiramate: a comparison of the impact of age and comedication. Ther Drug Monit. 2012;34(4):440–5.

31. Ahn JE, Birnbaum AK, Brundage RC. Inherent correlation between dose and clearance in therapeutic drug monitoring settings: possible misinterpretation in population pharmacokinetic analyses. J Pharmacokinet Pharmacodyn. 2005;32(5-6):703–18.

32. Perucca E, Gidal BE, Baltès E. Effects of antiepileptic comedication on levetiracetam pharmacokinetics: a pooled analysis of data from randomized adjunctive therapy trials. Epilepsy Res. 2003;53(1-2):47–56.

33. Coupez R, Nicolas J-M, Browne TR. Levetiracetam, a new antiepileptic agent: lack of in vitro and in vivo pharmacokinetic interaction with valproic acid. Epilepsia. 2003;44(2):171–8.

34. Snoeck E, Stockis A. Dose-response population analysis of levetiracetam add-on treatment in refractory epileptic patients with partial onset seizures. Epilepsy Res. 2007;73(3):284–91.

35. Silverstein FS, Ferriero DM. Off-label use of antiepileptic drugs for the treatment of neonatal seizures. Pediatr Neurol. 2008;39(2):77–9.

36. Agrawal A, Banergee A. A Review on Pharmacokinetics of Levetiracetam in Neonates. Curr Drug Metab. 2017 16;18(8):727–34.

37. Mruk AL, Garlitz KL, Leung NR. Levetiracetam in neonatal seizures: a review. J Pediatr Pharmacol Ther. 2015;20(2):76–89.

38. Perucca E. Clinical pharmacokinetics of new-generation antiepileptic drugs at the extremes of age. Clin Pharmacokinet. 2006;45(4):351–63.

39. Bansal R, Suri V, Chopra S, Aggarwal N, Sikka P, Saha SC, et al. Change in antiepileptic drug prescription patterns for pregnant women with epilepsy over the years: Impact on pregnancy and fetal outcomes. Indian J Pharmacol. 2019;51(2):93–7.

40. Daugaard CA, Sun Y, Dreier JW, Christensen J. Use of antiepileptic drugs in women of fertile age. Dan Med J. 2019;66(8).

41. Koc G, Keskin Guler S, Karadas O, Yoldas T, Gokcil Z. Fetal safety of levetiracetam use during pregnancy. Acta neurologica Belgica. 2018;118(3):503–8.

42. Voinescu PE, Pennell PB. Management of epilepsy during pregnancy. Expert Rev Neurother. 2015;15(10): 1171–87.

43. Garrity LC, Turner M, Standridge SM. Increased levetiracetam clearance associated with a breakthrough seizure in a pregnant patient receiving once/day extended-release levetiracetam. Pharmacotherapy. 2014;34(7):e128–32.

44. Thangaratinam S, Marlin N, Newton S, Weckesser A, Bagary M, Greenhill L, et al. AntiEpileptic drug Monitoring in PREgnancy (EMPiRE): a double-blind randomised trial on effectiveness and acceptability of monitoring strategies. Health Technol Assess. 2018;22(23):1–152.

45. Polepally AR, Pennell PB, Brundage RC, Stowe ZN, Newport DJ, Viguera AC, et al. Model-based lamotrigine clearance changes during pregnancy: clinical implication. Ann Clin Transl Neurol. 2014;1(2):99–106.

